# Circulating adipokines in non-obese PCOS patients: a systematic review and meta-analysis

**DOI:** 10.1101/2020.10.11.20210716

**Authors:** Kainan Lin, Xiaoting Sun, Xiao Wang, Hanchu Wang, Xia Chen

## Abstract

Concentrations of circulating adipokines in non-obese polycystic ovary syndrome (PCOS) patients had been reported in many researches, however, these results were conflicting. The aim of this meta-analysis was to assess whether the levels of circulating adipokines were changed in non-obese PCOS. To identify eligible studies, literature research was performed in the database of PubMed, Embase, Web of Science without the restriction of region, publication or language. Of the total studies found, only 81 met the inclusion criteria. The meta-analysis showed that circulating levels of adiponectin [-0.95 (95% CI, −1.36 to −0.53)] decreased statistically in non-obese PCOS women. On the contrary, circulating levels of chemerin [1.13 (95% CI, 0.08 to 2.18)], leptin [0.47 (95% CI, 0.13 to 0.81)], resistin [0.45 (95% CI, 0.03 to 0.88)] and visfatin [1.38 (95% CI, 0.68 to 2.09)] increased significantly in non-obese PCOS females. Besides, there was no statistically significant change in the circulating levels of apelin [0.32 (95% CI, −1.34 to 1.99), irisin [1.01(95% CI, −0.68 to 2.70), omentin [-0.37(95% CI, −1.05 to 0.31)] and vaspin [0.09(95% CI, −0.14 to 0.32)] in non-obese PCOS patients. Scientific evidence suggested that the levels of circulating adipokines altered in non-obese PCOS patients compared with controls. Independent of the degree of obesity, the abnormal change of circulating adipokines levels might play an important role in the occurrence and development of PCOS.

## 1. Introduction

PCOS is a common endocrine and metabolic disease, which affects nearly affects 6%-10% of reproductive-aged women according different criteria^1^. PCOS is diagnosed by sparse ovulation or anovulation, hyperandrogenism or/and polycystic ovaries^2^. Current studies have proved that the obesity rate of PCOS patients is significantly increased, which suggests that obesity may be related to the occurrence and symptoms of PCOS^3^.

Fat tissue, which we often think of as a storage site for energy, is a crucial endocrine tissue in the body^4^. Adipokine is an active hormone and factor secreted by adipocytes, including adiponectin, leptin, omentin, etc. These hormones and factors secreted by adipose tissue are involved in many critical physiological processes in the body. Therefore, abnormal changes in adipokines may also lead to some endocrine diseases in the body^5^. Besides, studies have shown that adipokines play an essential role in the pathogenesis of obesity and obesity-related diseases^6^.

PCOS patients are likely to have metabolic complications such as type 2 diabetes, IR (insulin resistance), and adipose tissue dysfunction ^7-9^. While IR affects about 10% to 25% of the general population, compared with two to three times the risk in PCOS patients ^10, 11^. The incidence of obesity is increased in patients with PCOS [RR (95 CI): 2.77 (1.88, 4.10)] compared with patients without PCOS^12^. And the symptoms in PCOS patients who combined with obesity aggravates significantly. However, up to now, the mechanism of PCOS has not been determined.

Dysfunction of adipose tissue can lead to changes in adipokine levels^13^. Many studies have proved that there are changes in adipokines levels in obese PCOS patients, indicating that adipokines play a role in obese PCOS patients^14, 15^. But so far, studies on adipokines levels in non-obese PCOS patients have been inconsistent. Moreover, it is not known whether the changes of adipokines are related to PCOS directly or obesity or both. Thus, we aimed to perform this systematic review and meta-analysis to evaluate the levels of different kinds of adipokines in non-obese PCOS women.

## 2. Materials and Methods

### 2.1. Search Strategy

This systematic review and meta-analysis were designed according to the Preferred Reporting Items for Systematic Reviews and Meta-Analyses (PRISMA) statement^16^ and Meta-analysis Of Observational Studies in Epidemiology (MOOSE)^17^.

### 2.2. Eligibility Criteria

Studies defining PCOS conforming to the Rotterdam Criteria or other compatible criteria were included. (2) Studies designed about obese women with PCOS were excluded from the study. (3) Reviews, non-human studied and conference proceedings were excluded. (4) Studies without control groups were also excluded. (5)

Studies without extractable data(not provided as mean±SD) were excluded.

### 2.3. Information Sources

To identify eligible studies, exhaustive literature was performed in the electronic database of PubMed, Embase, Web of Science without the restriction of region, publication or language. All articles published before July 2019 was considered for eligibility.

### 2.4. Search Terms

The search strategy involved the identification of all possible studies using combinations of the following keywords:(“adiponectin,” or “apM-1 Protein,” or “ACRP30 Protein,” ; “apelin,” ; “chemerin,” or “TIG2 protein,” ; “irisin,” or “FRCP2 protein,” ; “leptin,” or “Obese Protein,” ; “omentin,” or “intelectin 1,” ; “resistin,” ; “vaspin,” or “SERPINA12 protein,”; “visfatin,” or “NAMPT Protein,”) and (“Polycystic Ovary Syndrome,” “PCOS,”). Data on the levels of adiponectin, apelin, chemerin, irisin, leptin, omentin, resistin, vaspin, visfatin was extracted.

### 2.5. Data Extraction

Two reviewers reviewed all the literature searched and determined whether they met the inclusion criteria independently. Discrepant opinions between the two reviewers were resolved by discussion and consultation with a third reviewer, if necessary. The following information will be obtained from the literature: General study characteristics (name of the first author, year of publication, study location), Age and BMI (body mass index) of participants and summary of conclusions of the study. We used the available data for our analysis.

### 2.6. Quality Assessment

We used Cochrane Collaboration’s tool to assess the quality of studies included. Seven domains were evaluated by this tool, including random sequence generation, allocation concealment, blinding of participants and personnel, outcome assessment, incomplete outcome data, selective reporting, and other biases. Each indicator has three levels: high risk, low risk and unknown risk. Each indicator was evaluated separately by two reviewers and the differences encountered in the process were unified by discussion. The possibility of publication bias was assessed by visual inspection of the funnel plot and egger’s test.

### 2.7. Statistical Analysis

The software of Review Manager Version 5.3 was used to perform the effects by meta-analysis and to construct forest plot. The expected outcome of each study was a difference in average adipokines levels between PCOS patients and healthy controls. Due to the different measurement methods for each study, Review Manager was used to calculate the standard mean difference (SMD) with 95% confidence intervals. The Cochran’s chi-square-based Q statistic test and the I2 test were calculated to assess potential heterogeneity between the individual studies. According to significant heterogeneity, moderate heterogeneity and low heterogeneity, we selected a random effect model and a fixed effect model respectively. STATA15.0 version was used for egger’s test to check publication bias and trim-and-fill method. The t value was used to determine whether there was publication bias. When t value ≥0.05, it was considered that there might be no publication bias.

## 3. Result

### 3.1. Study Selection

Our search strategy identified 1540 potential articles. One thousand two hundred fifty-six studies were excluded after screening based on title or abstract, and 284 potentially relevant studies were assessed by reviewing the full-text article. Among these studies, 203 articles were excluded from the meta-analysis and systematic review owing to lack of control groups or obese subjects. Since data in some articles did not present as Mean ± SD, we continued to exclude 10 articles. Finally, 71 studies including 2495 subjects with PCOS and 2520 controls met our inclusion criteria for the meta-analysis. Figure 1 presents the search strategy for study selection.

**Fig 1.**
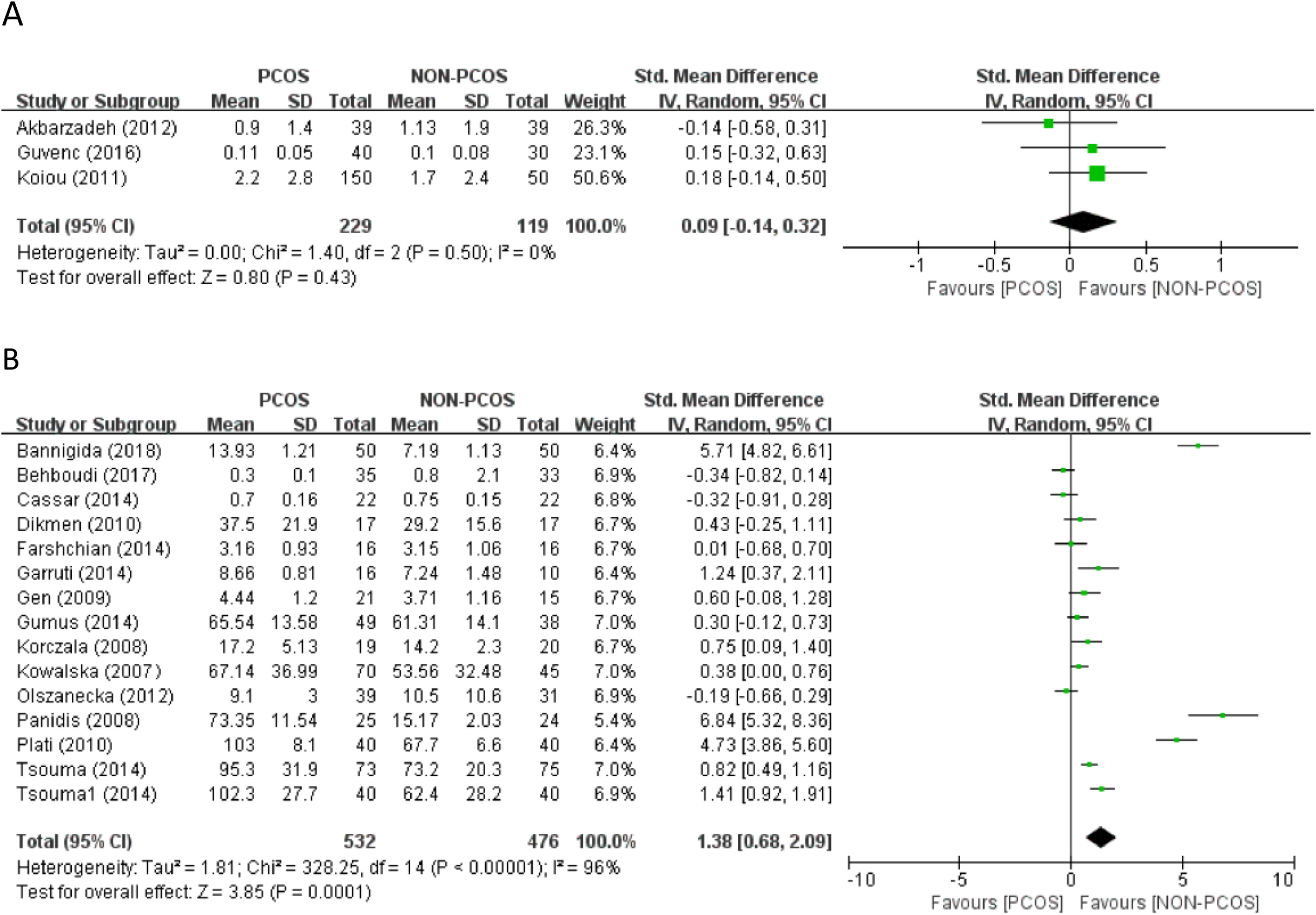
Flow chart of study inclusion in systematic review and meta-analysis.

### 3.2. Characteristics of Included Studies

The features of the included literature were presented in Table1. Among the 81 included studies, there are several diagnostic criteria for PCOS: The Rotterdam Criteria were adopted in fifty-three articles; The ESHRE/ASRM consensus had been used in three articles; Nine of the articles used the NIH Criteria; Two article used the Criteria of National Institute of Child Health and Human Disease; Other articles adopted some of the specific standards mentioned in the article. The included studies covered twenty-six countries :There were eight articles from China; Two from Saudi Arabia; nineteen studies from Turkey; One study from Croatia; One article from Netherlands; one article from Pakistan; One article from Denmark; Three articles from India; Four studies from Iran; Three studies from South Korea; Three articles from Taiwan; One studies were based in France; One article was based in the United States; Four articles were based on research in Egypt; seven articles on Greece; Four articles from Italy; one article from Israel; one article from Indonesia; Eight articles were written in Poland; one article from Iraq; In addition to one article in Germany; Brazil contributed two articles; an article from Japan; one article from Australia; eventually there were one articles from Spain and one article from Qatar. The subjects included in the study ranged in age from youth to old age. One criterion for obesity was BMI greater than or equal to 30.0, so the studies we included had a BMI lower than 30.0. The Table1 also briefly listed the primary conclusions of each article.

**Table 1.**
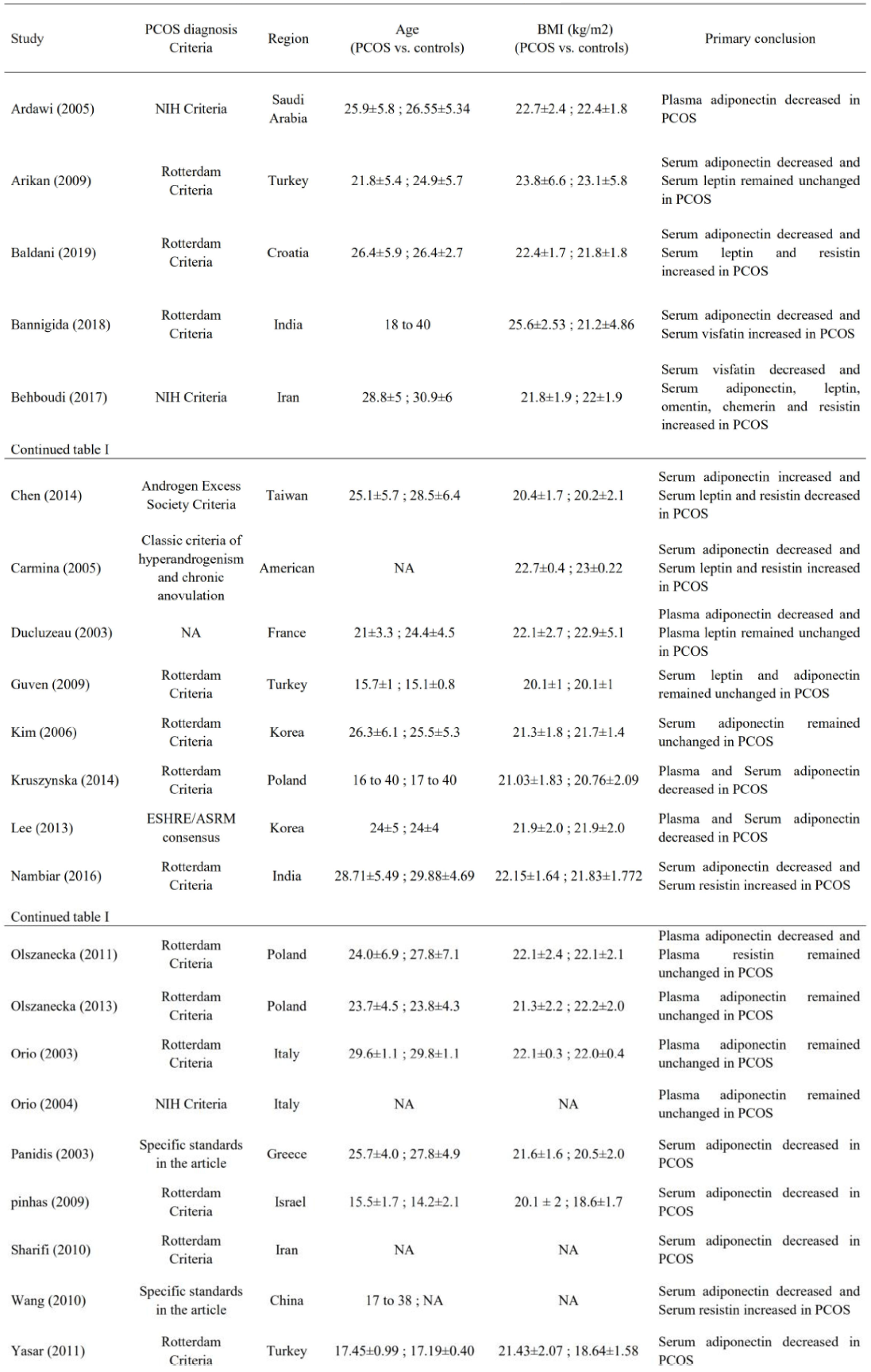
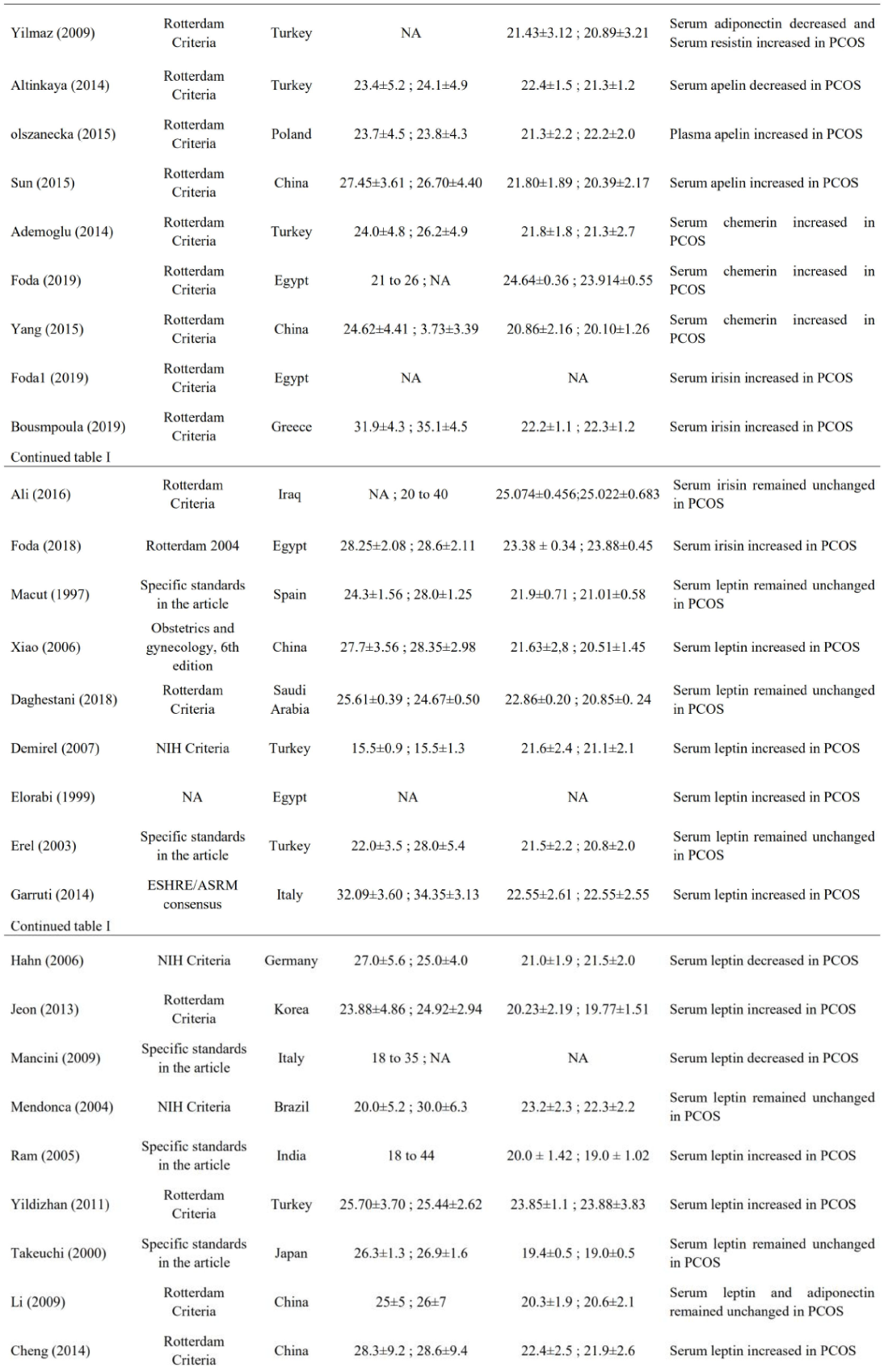
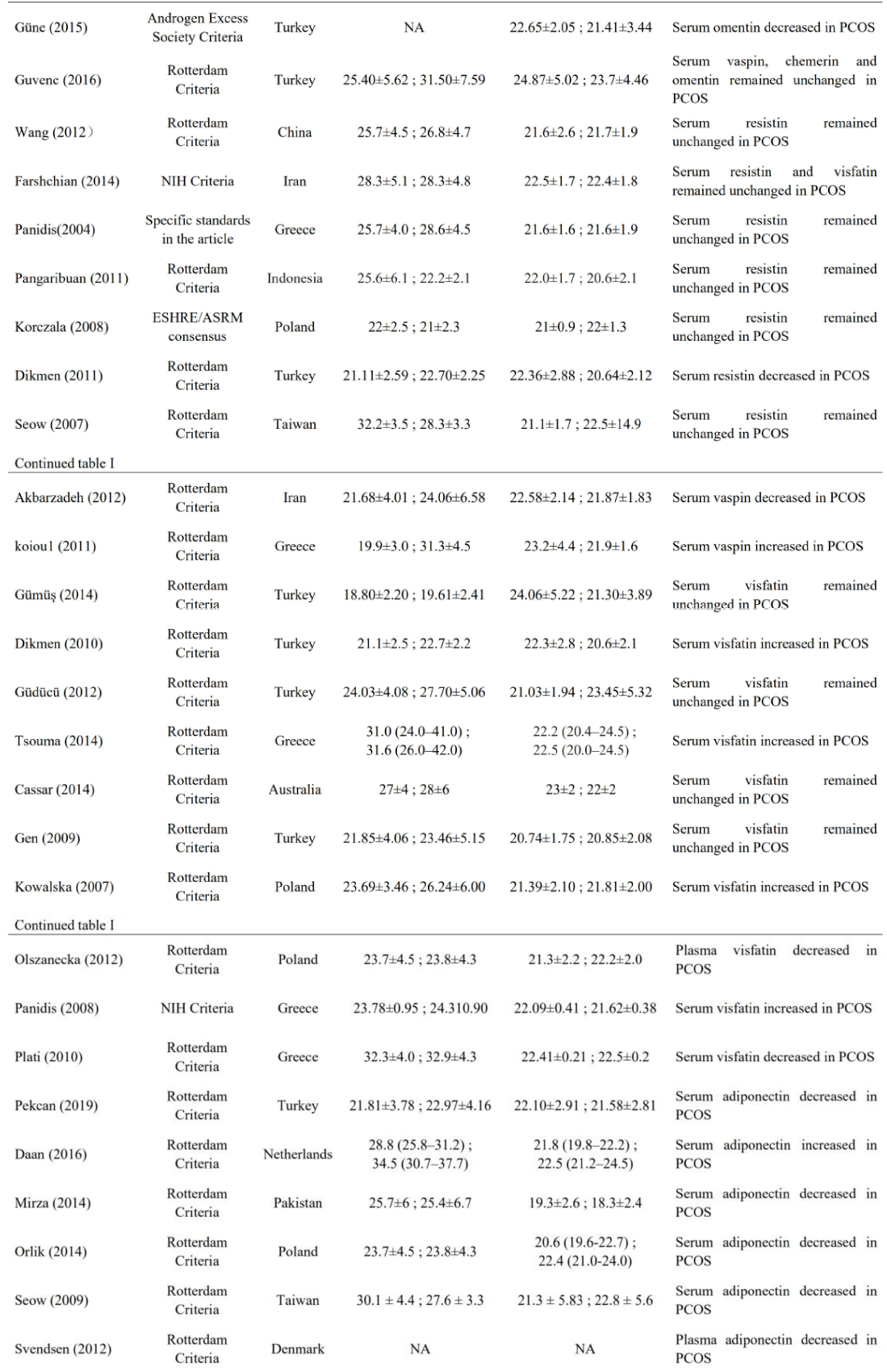
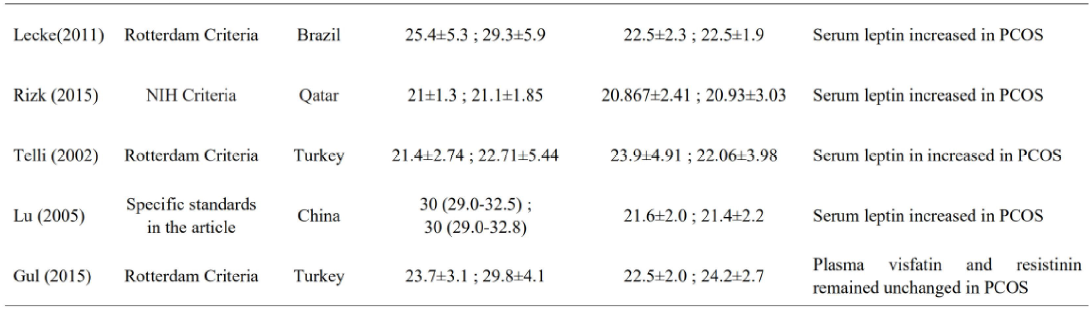
Characteristic of individual study included in the systematic review and meta-analysis. Abbreviations: PCOS, polycystic ovary syndrome; BMI, body mass index; NA, not available; NIH, National Institutes of Health.

### 3.3. The relationship between different kinds of adipokines levels and PCOS

Thirty studies reported the level of adiponectin in non-obese PCOS women as illustrated in Figure 2A which showed the forest plot of those studies; there was significant heterogeneity among studies (I-squared = 95%, *p*<0.00001). Overall, PCOS was associated with a change in adiponectin level of −0.95 (95% CI, −1.36 to −0.53; *P* < 0.00001; n = 30 studies, 2565 participants).

**Fig 2.**
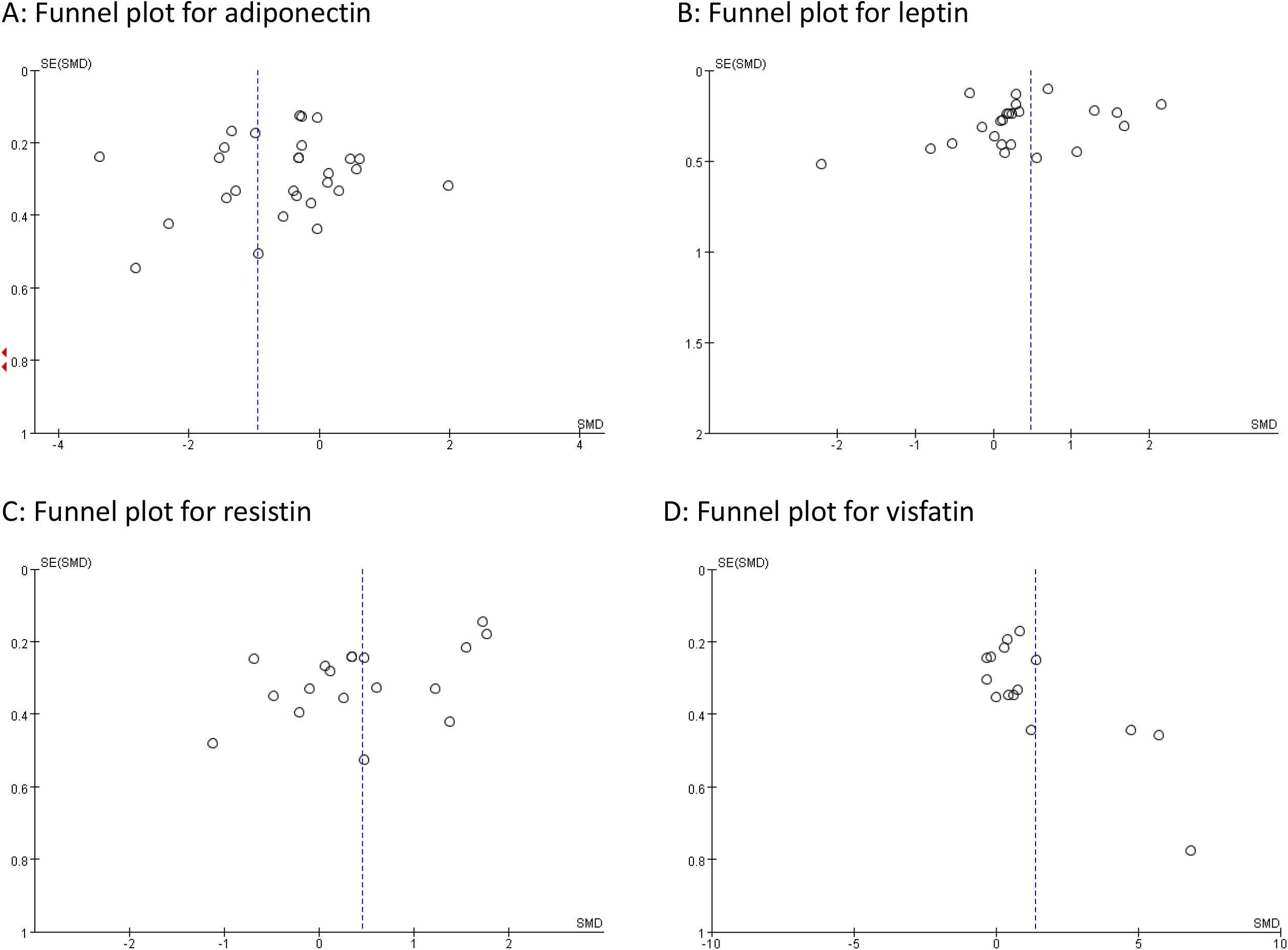
Meta-analysis of the association between the levels of adipokines in non-obese patient and PCOS. Weights are from random effects analysis. A: Meta-analysis of adiponectin. B: Meta-analysis of apelin. C: Meta-analysis of chemerin. D: Meta-analysis of irisin. Abbreviations: PCOS, polycystic ovary syndrome; CI, confidence interval; SD, standard difference.

The four trials (n = 254 participants) focused on levels of apelin in non-obese PCOS patients. However, no significant difference was noted between PCOS and circulating apelin [0.32 (95% CI, −1.34 to 1.99; *P* = 0.70]. The forest plot of those studies was shown in Figure 2B.

Five individual studies were included to compare the levels of chemerin between PCOS and controls. The meta-analysis showed an increased level of 1.13 (95% CI = 0.08 to 2.18; *P* = 0.03; Figure 2C). There was significant heterogeneity among these studies (*I*^2^ = 96%).

Four studies reported the level of irisin in non-obese PCOS patients as illustrated in Figure 2D which showed the forest plot of those studies; there was significant heterogeneity among studies (I-squared = 97%, *P*<0.00001). Overall, PCOS show no correlation with circulating irisin levels of 1.01 (95% CI, −0.68 to 2.70; *P* = 0.24; n = 4 studies, 282 participants).

The twenty-five trials (n = 2148 participants) focusing on levels of leptin in non-obese PCOS patients found a statistically increase level of 0.47 (95% CI, 0.13 to 0.81; *P* = 0.007). The forest plot of those studies was showed in Figure 3A.

**Fig 3.**
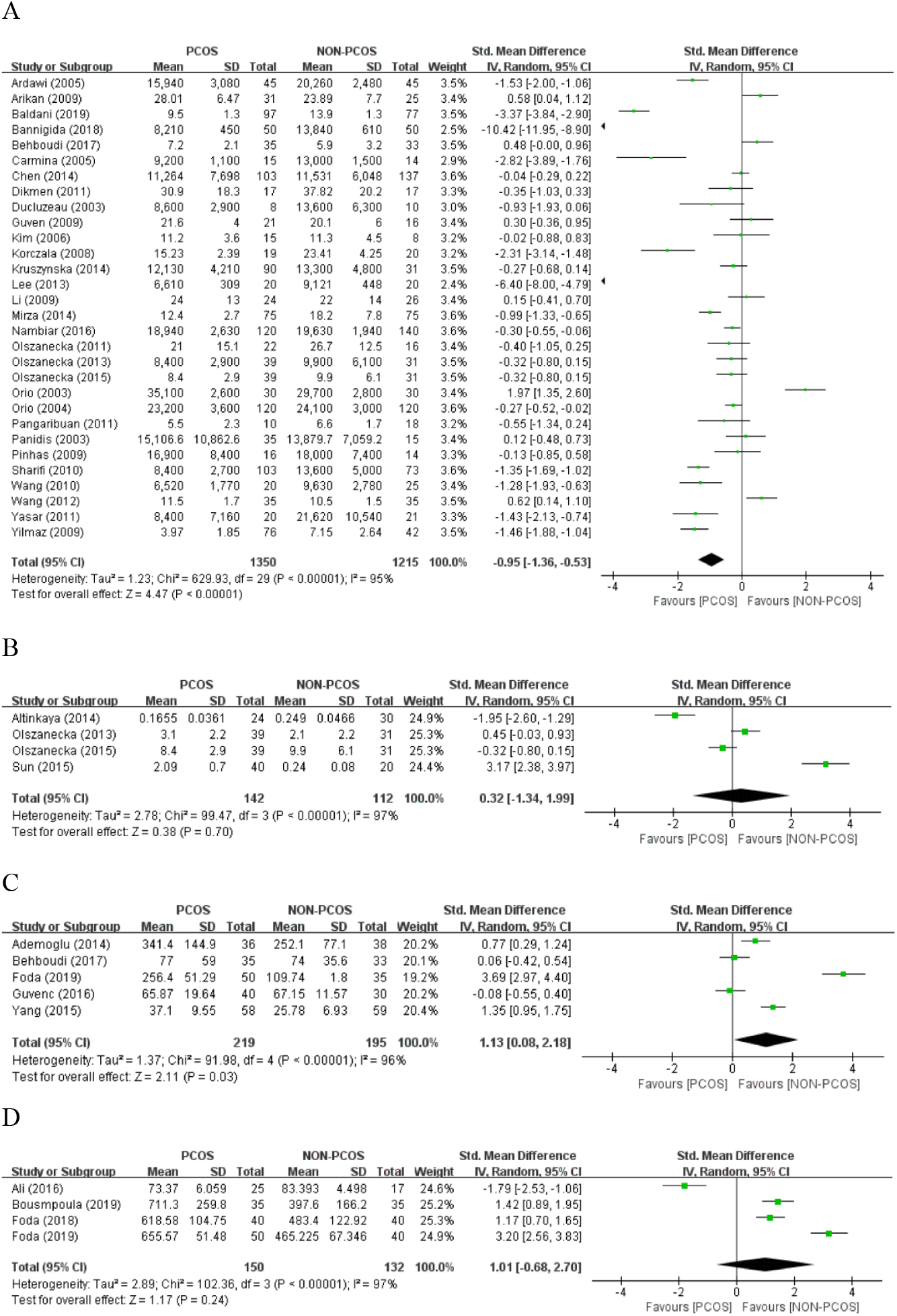
Meta-analysis of the association between the levels of adipokines in non-obese patient and PCOS. Weights are from random effects analysis. A: Meta-analysis of leptin. B: Meta-analysis of omentin. C: Meta-analysis of resistin. Abbreviations: PCOS, polycystic ovary syndrome; CI, confidence interval; SD, standard difference.

Five individual studies were included to compare the levels of omentin between non-obese PCOS patients and controls. The meta-analysis showed the equal level of omentin [-0.37 (95% CI = −1.05 to 0.31; *P* = 0.29; Figure 3B)]. There was significant heterogeneity among these studies (*I*^2^ = 89%).

Eighteen studies reported the level of resistin in non-obese PCOS women as illustrated in Figure 3C which showed the forest plot of those studies; there was significant heterogeneity among studies (I-squared = 91%, *P*<0.00001). Overall, PCOS was associated with an increase in resistin level of 0.45 (95% CI, 0.03 to 0.88; *P* = 0.03; n = 28 studies, 1223 participants).

The three trials (n = 348 participants) focusing on levels of vaspin in patients with PCOS found a nonsignificant change level of 0.09 (95% CI, −0.14 to 0.32; *P* = 0.43). The forest plots of those studies were showed in Figure 4A.

**Fig 4.**
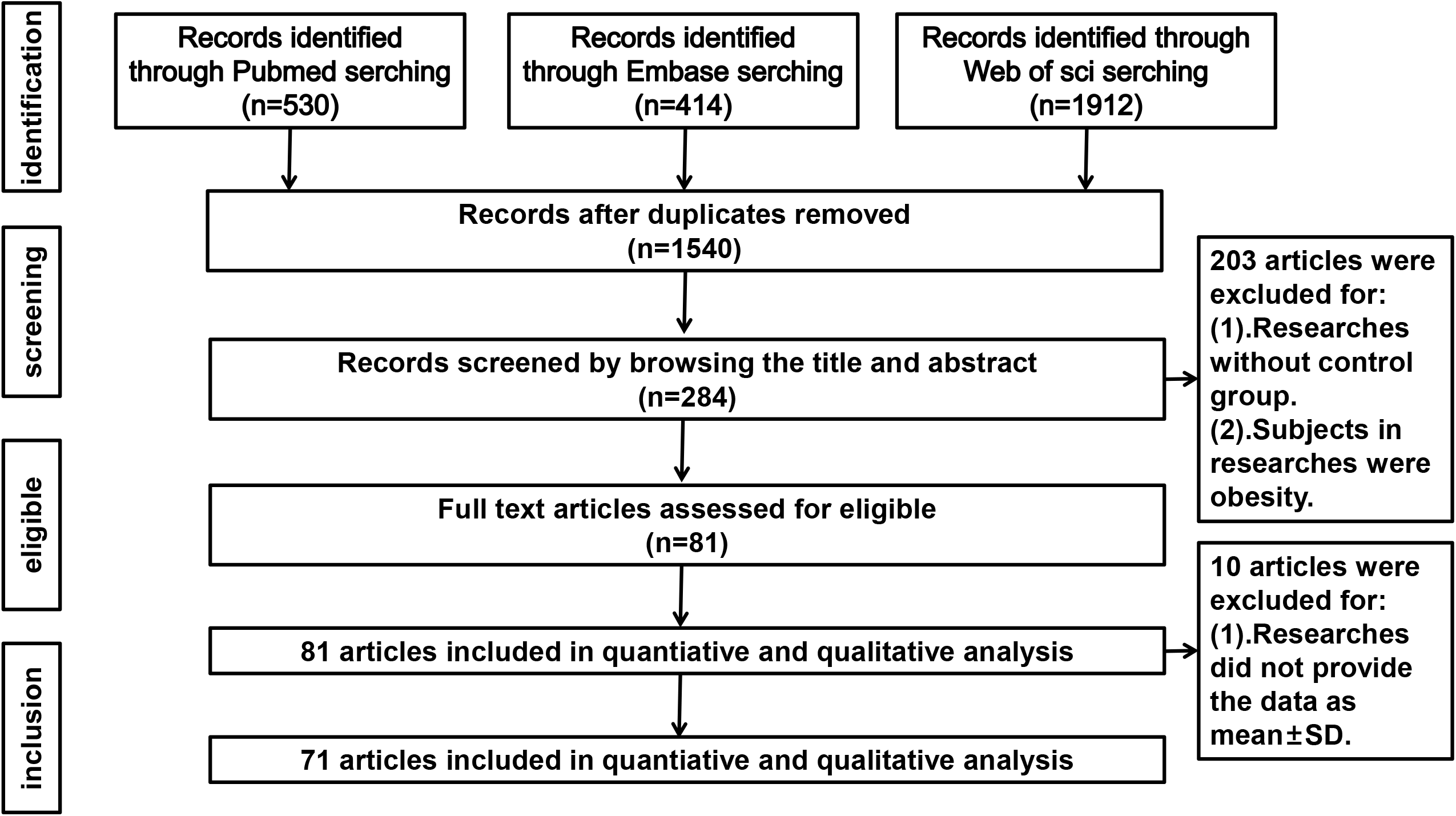
Meta-analysis of the association between the levels of adipokines in non-obese patient and PCOS. Weights are from random effects analysis. A: Meta-analysis of vaspin. B: Meta-analysis of visfatin. Abbreviations: PCOS, polycystic ovary syndrome; CI, confidence interval; SD, standard difference.

Twenty-six individual studies were included to compare the level of visfatin between PCOS group and controls. The meta-analysis showed an increased level of 1.38 (95% CI = 0.68 - 2.09; *P* = 0.0001; Figure 4B). There was significant heterogeneity among these studies (*I*^2^ = 96%).

### 3.4. Publication Bias

Funnel plot method was used to detect publication bias and Egger’s test was used to quantify publication bias. The shape of the funnel plot did not reveal any obvious asymmetry in adiponectin, resistin. However, it seemed that the funnel plot of leptin and visfatin showed publication bias observably (Figure5A.5B.5C.5D). Egger’s test was used to provide statistical evidence of funnel plot symmetry. The results still did not suggest any evidence of publication bias in adiponectin and leptin. Egger’s test showed there was possible publication bias among studies in resistin and visfatin (Table 2). We used the trim-and-fill method to recalculate our pooled risk estimate of resistin and visfatin. Meta-trim showed that corrected SMD (standard mean difference) was not differ from uncorrected SMD, which suggested that publication bias had a small effect on the final result. We did not assess the publication bias for apelin, chemerin, irisin, omentin and vaspin based on the Cochrane Handbook for Systematic Reviews of Interventions (www.cochranehandbook.org), which stated that the test for publication bias yields unreliable results when less than ten studies were included in a meta-analysis.

**Fig 5.**
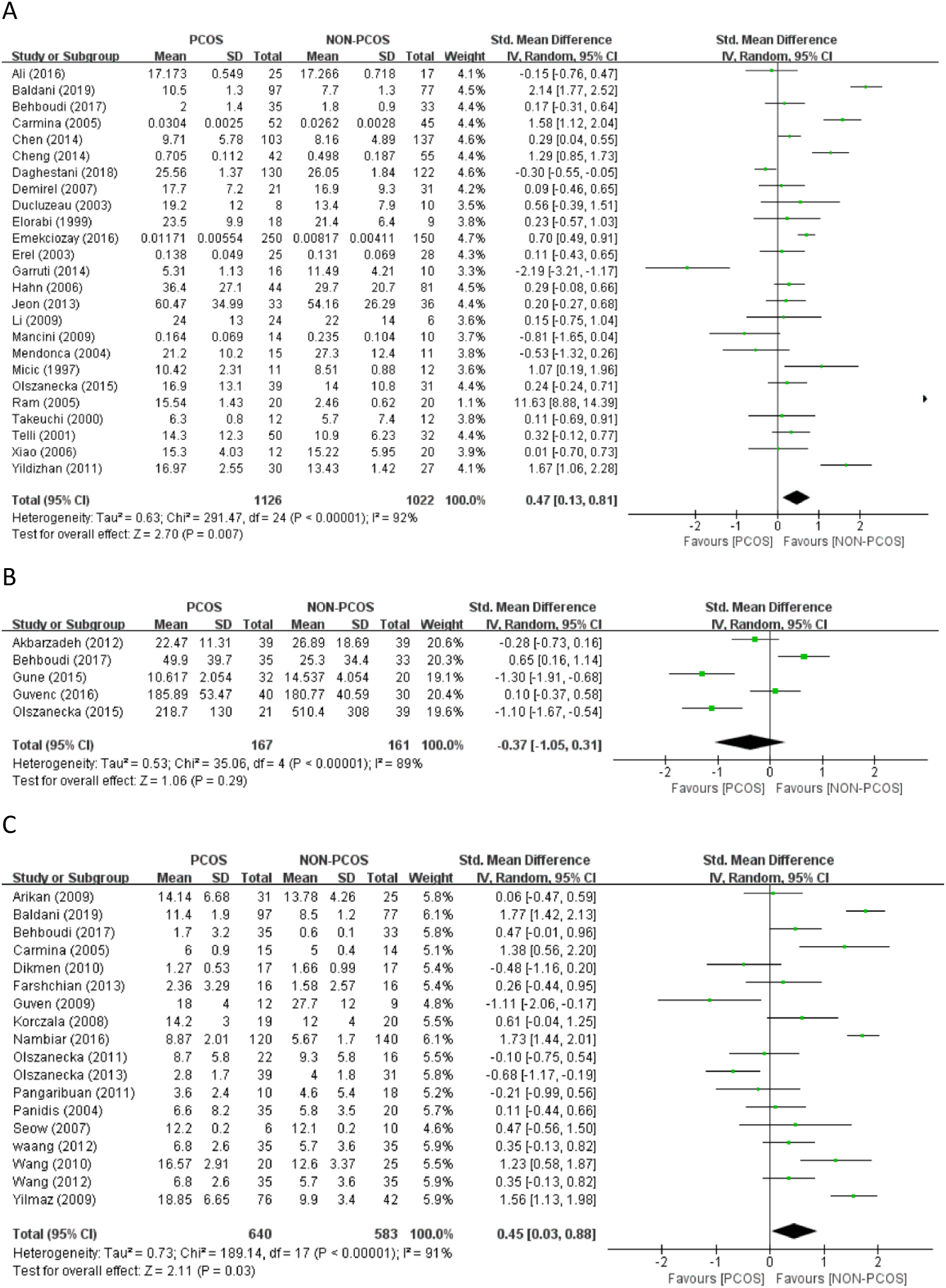
Funnel plot analysis to detect publication bias under a dominant model. A: Funnel plot for adiponectin; B: Funnel plot for leptin; C: Funnel plot for resistin; D: Funnel plot for visfatin. Abbreviations: SE, standard error; SMD, standard mean difference.

**Table 2.** Egger publication bias test for the adiponectin, leptin, visfatin and resistin. Abbreviations: CI, confidence interval.

| Indices | No. of studies | Coefficient | Standard Error | t | P | 95%CI |
| --- | --- | --- | --- | --- | --- | --- |
| adiponectin | 30 | -3.45 | 1.98 | 1.75 | 0.092 | -7.50 to 0.60 |
| leptin | 25 | 0.45 | 1.54 | 0.29 | 0.775 | -2.75 to 3.64 |
| visfatin | 15 | 8.39 | 3.11 | 2.7 | 0.018 | 1.67 to 15.11 |
| resistin | 18 | -6.48 | 1.95 | 3.33 | 0.004 | -10.61 to -2.35 |

## 4. Discussion

To our knowledge, this was the first systematic review and meta-analysis aimed at quantifying the levels of nine kinds of adipokines in non-obese PCOS patients. This systematic review and meta-analysis, including 81 studies demonstrated that non-obese PCOS women showed a decreasing level of adiponectin and the increased levels of leptin, visfatin, chemerin and resistin respectively. Our results indicated that the concentration of common adipokines in plasma or serum changed remarkably in non-obese PCOS women.

Over the past decade, a large number of studies had investigated the levels of circulating adipokines in non-obese PCOS patients. Since the levels of adipokines differed from the geographic regions, ethnicity or age, the results of studies were inconsistent. For example, one Mahmoud A’s paper showed lower levels of adipokines in PCOS patients than in the control group^18^ whereas an Arikan’s paper showed the opposite result^19^. The purpose of meta-analysis was to integrate these results for statistical analysis. Combining the findings of these studies was an attractive option to enhance the credibility of the evidence^20^. Therefore, we performed a meta-analysis to reach a reliable conclusion on the changes of adipokines concentration in non-obese PCOS patients.

Considering that studies had shown the levels of adipokines was related to BMI ^21^ and there was evidence that PCOS patients had an increased BMI^22^, so the change in adipokines levels might be caused by obesity rather than the PCOS itself. In this meta-analysis, we only included the study on non-obese PCOS patients, so we could exclude the influence of obesity and directly analyzed the relationship between PCOS and adipokine levels.

Understanding the disturbance of adipokines levels in non-obese PCOS patients help to explain the pathophysiology and symptoms of PCOS:(1). Hyperandrogenism: the occurrence of PCOS was closely related to hyperandrogenism^23^. Animal studies had shown that impaired insulin sensitivity and glucose intolerance in mice (selectively knocked down androgen receptors in adipocytes) were not related to obesity. Besides, it was related to the changes of adipokines levels. Therefore, it could be presumed that the AR (androgen receptor) in fat cells can control insulin sensitivity and glucose tolerance independently of obesity and affect adipokines levels in the meanwhile^24^. In non-obese PCOS patients, androgen hypersecretion and androgen receptor dysfunction were the main symptoms, which might lead to the changes in adipokines levels^25^. Meanwhile, some studies had shown that gender had a certain influence on the levels of adipokines in the body, so the increase of androgens in PCOS patients might lead to the change of the levels of adipokines in the body^26^. (2). IR: there was the evidence that the occurrence of PCOS was closely related to IR^27^. In vitro studies had shown that in vitro fat cells, certain adipokines activate protein kinases that increased insulin-mediated glucose transport, thus increasing insulin sensitivity through specific pathways, such as omentin^28^. Meanwhile, some studies had shown the expression of proinsulin mRNA was inhibited by leptin in high level, which led to the decreased transcription activity of insulin gene promoter and the inhibited phosphorylation of insulin receptor matrix in peripheral tissues^29, 30^. These studies suggested that IR might affect adipokines levels by increasing insulin levels. High leptin level could lead to IR in non-obese PCOS patients in the following possible ways: inhibition of phosphoenolpyruvate carboxykinase inhibited hepatic glucose oxidation and increased hepatic glycogen reserve; deposit of fat in skeletal muscle cells; breakdown of fat and the production of FFA (free fatty acids); direct inhibition of basal insulin secretion and glucose-stimulated insulin secretion. In addition, the insulin sensitizer thiazolidinedione could up-regulate the expression of adiponectin gene and promote the differentiation and apoptosis of adipocytes, thus reducing IR. Therefore, we inferred that disorders of adipokines levels played an important role in IR of non-obese PCOS patients. We might be able to treat PCOS with drugs that correct the disorder of adipokines levels. (3). Chronic inflammation: Chemerin, identified as an inflammatory factor at first, was thought to promote chemotaxis of immature dendritic cells and macrophages through its receptor CMKLR1^31^. CMKLR1 had been found to be expressed in many immune cells, including immature dendritic cells, myeloid dendritic cells, macrophages and natural killer cells^32, 33^. This meant that chemerin and its receptor CMKLR1 were involved in the recruitment of different immune cells to the site of injury, and might affect the occurrence and development of inflammation. Adipokines and their receptors were elevated in many inflammatory states, suggesting that adipokines levels might be predictors of PCOS. Omentin had anti-inflammatory, anti-cardiovascular and anti-diabetic effects. Considering that obesity was a chronic low-grade inflammatory disease, long-term inflammatory stimulation might be one of the reasons for omentin’s down-regulated expression. ^34, 35^.

In the long run, non-obese PCOS patients with disturbance of adipokine levels might lead to more related diseases, such as hypertension, coronary heart disease, type 2 diabetes, etc^36^. Type 2 diabetes was almost 10 times more common in PCOS patients than in the normal population, and glucose intolerance was 30% to 50% higher in obese PCOS patients^37^. PCOS in the future risk of cardiovascular disease (CVD) should not be ignored. PCOS was also closely related to IR, which could lead to a variety of cardiac metabolic abnormalities (such as dyslipidemia, high blood pressure, glucose intolerance, diabetes and metabolic syndrome) that increased the risk of cardiovascular disease in women^38^.

Therefore, adipokines levels were of great significance in the diagnosis and treatment of PCOS, and people should improve their understanding of abnormal changes in adipokines levels. Metformin might alleviate corresponding symptoms and treat PCOS by correcting abnormal levels of certain adipokines. For example, it could reduce the levels of resistin, visfatin, irisin, chemerin and so on^39-42^. A recent report suggested that insulin and glucose affected the secretion of visfatin through phosphatidylinositol 3-kinase and protein kinase B pathway^43^. Metformin could increase the sensitivity of peripheral tissues to insulin which help to reduce the hyperinsulinemia caused by insulin resistance in PCOS patients. In the above ways, metformin could alleviate the abnormal change of visfatin level. Moreover, adipokines and hyperandrogenemia formed a vicious cycle of endocrine metabolism, promoting the risk of PCOS and other related endocrine diseases. Metformin could also relieve the symptoms of PCOS patients by reducing their high testosterone levels through specific mechanisms^44^. We presumed that metformin might be used to correct the disorder of adipokine levels and thus to treat PCOS, but more experiments are needed to prove.

Statistically, significant heterogeneity was found in the analysis of adiponectin, apelin, chemerin, irisin, leptin, omentin, resistin, visfatin might reflect the clinical heterogeneity and PCOS diagnostic criteria, different ethnic backgrounds, national setting, age, or BMI. In addition, the BMI to determine obesity was different. This suggested that caution should be exercised in extrapolating these results to more extensive applications. However, the analysis of apelin showed no heterogeneity, indicating that the included pieces of literature were homogeneous, and the study results were accurate. Second, we found evidence of publication bias. Although a trim-and-fill method suggested that unpublished trials might not influence the levels of resistin and visfatin in non-obese patients with PCOS, these results should be interpreted carefully.

Our literature search was comprehensive, and we did not apply any restrictions on language to limit our ability to assess the relationship between the levels of adipokines and PCOS. However, our meta-analysis still had some limitations. First of all, there were many different test methods for the identification of PCOS, so the diagnose of PCOS had heterogeneity. In addition, changes in adipokines levels in normal weight or overweight PCOS patients were studied in this study, but we could not entirely exclude all obese participants in the included study due to the different criteria. Finally, there were many different methods to determine the levels of adipokine in the blood, which may lead to some deviation. Despite these limitations, the present meta-analysis had increased the statistical power by pooling the results of single studies. Therefore, the total number of subjects was sufficiently large to support our conclusion.

In summary, this systematic review and meta-analysis demonstrated that the levels of circulating adipokines in non-obese PCOS patients with was changed to varying degrees, which help to find potential pathogenesis and new biochemical diagnostic criteria of PCOS. Moreover, this result might provide a new scheme to treat PCOS by correcting disturbance of adipokines levels. However, further studies were needed to explore the potential mechanism of the disturbance and focus on whether the PCOS could be treated by correcting the levels of adipokines.

## Data Availability

All data, models, and code generated or used during the study appear in the submitted article

## Author’s roles

K.L. conducted literature search, complied data and drafted manuscript. X.S. and X.W. contributed to literature search and data interpretation. H.W. reviewed manuscript and provided advice. C.X contributed to critical discussion, reviewed all drafts of this article, provided extensive advice, and revised the manuscript.

## References

1. Bozdag G, Mumusoglu S, Zengin D, et al. The prevalence and phenotypic features of polycystic ovary syndrome: a systematic review and meta-analysis. Hum Reprod 2016; 31: 2841–2855. DOI: 10.1093/humrep/dew218.

2. Palomba S, de Wilde MA, Falbo A, et al. Pregnancy complications in women with polycystic ovary syndrome. Human reproduction update 2015; 21: 575-592. 2015/06/29. DOI: 10.1093/humupd/dmv029.

3. Joham AE, Palomba S and Hart R. Polycystic Ovary Syndrome, Obesity, and Pregnancy. Semin Reprod Med 2016; 34: 93–101. DOI: 10.1055/s-0035-1571195.

4. Holst D and Grimaldi PA. New factors in the regulation of adipose differentiation and metabolism. Current opinion in lipidology 2002; 13: 241–245.

5. Havel PJ. Control of energy homeostasis and insulin action by adipocyte hormones: leptin, acylation stimulating protein, and adiponectin. Current opinion in lipidology 2002; 13: 51–59.

6. Chen T, Wang F, Chu Z, et al. Serum CTRP3 Levels In Obese Children: A Potential Protective Adipokine Of Obesity, Insulin Sensitivity And Pancreatic β Cell Function. Diabetes, metabolic syndrome and obesity : targets and therapy 2019; 12: 1923–1930. DOI: 10.2147/dmso.S222066.

7. Legro RS, Kunselman AR, Dodson WC, et al. Prevalence and predictors of risk for type 2 diabetes mellitus and impaired glucose tolerance in polycystic ovary syndrome: A prospective, controlled study in 254 affected women. Journal of Clinical Endocrinology & Metabolism 1999; 84: 165–169. DOI: 10.1210/jc.84.1.165.

8. Stepto NK, Cassar S, Joham AE, et al. Women with polycystic ovary syndrome have intrinsic insulin resistance on euglycaemic-hyperinsulaemic clamp. Human Reproduction 2013; 28: 777–784. DOI: 10.1093/humrep/des463.

9. Spritzer PM, Lecke SB, Satler F, et al. Adipose tissue dysfunction, adipokines, and low-grade chronic inflammation in polycystic ovary syndrome. Reproduction (Cambridge, England) 2015; 149: R219–R227. DOI: 10.1530/rep-14-0435.

10. Sirmans SM and Pate KA. Epidemiology, diagnosis, and management of polycystic ovary syndrome. Clinical Epidemiology 2014; 6: 1–13. DOI: 10.2147/clep.S37559.

11. Type 2 diabetes, polycystic ovary syndrome and the insulin resistance syndrome in adolescents: Are they one big iceberg? Paediatrics and Child Health 2002. DOI: 10.1093/pch/7.5.333.

12. Lim SS, Davies MJ, Norman RJ, et al. Overweight, obesity and central obesity in women with polycystic ovary syndrome: a systematic review and meta-analysis. Human reproduction update 2012; 18: 618–637. DOI: 10.1093/humupd/dms030.

13. Maury E and Brichard SM. Adipokine dysregulation, adipose tissue inflammation and metabolic syndrome. Molecular and cellular endocrinology 2010; 314: 1–16. DOI: 10.1016/j.mce.2009.07.031.

14. Itoh H, Kawano Y, Furukawa Y, et al. The role of serum adiponectin levels in women with polycystic ovarian syndrome. Clinical and experimental obstetrics & gynecology 2013; 40: 531–535.

15. Kumawat M, Ram M, Agarwal S, et al. Role of serum Leptin, insulin and other hormones in women with Polycystic ovarian syndrome. Indian Journal of Clinical Biochemistry 2018; 33: S88–S89. Conference Abstract. DOI: 10.1007/s12291-018-0795-1.

16. Knobloch K, Yoon U and Vogt PMJREDNHYD. Preferred reporting items for systematic reviews and meta-analyses (PRISMA) statement and publication bias. 2009; 18: e123.

17. Stroup DF, Berlin JA, Morton SC, et al. Meta-analysis of observational studies in epidemiology: a proposal for reporting. Meta-analysis Of Observational Studies in Epidemiology (MOOSE) group. Jama 2000; 283: 2008–2012. DOI: 10.1001/jama.283.15.2008.

18. Alfaqih MA, Khader YS, Al-Dwairi AN, et al. Correction: Lower Levels of Serum Adiponectin and the T Allele of rs1501299 of the ADIPOQ Gene Are Protective against Polycystic Ovarian Syndrome in Jordan. Korean journal of family medicine 2018; 39: 207. 2018/05/24. DOI: 10.4082/kjfm.2018.39.3.207.

19. Arikan S, Bahceci M, Tuzcu A, et al. Serum resistin and adiponectin levels in young non-obese women with polycystic ovary syndrome. Gynecological Endocrinology 2010; 26: 161–166. DOI: 10.3109/09513590903247816.

20. DerSimonian R and Laird N. Meta-analysis in clinical trials. Controlled clinical trials 1986; 7: 177–188.

21. Saremi A, Asghari M and Ghorbani A. Effects of aerobic training on serum omentin-1 and cardiometabolic risk factors in overweight and obese men. Journal of sports sciences 2010; 28: 993–998. DOI: 10.1080/02640414.2010.484070.

22. Barrea L, Arnone A, Annunziata G, et al. Adherence to the Mediterranean Diet, Dietary Patterns and Body Composition in Women with Polycystic Ovary Syndrome (PCOS). Nutrients 2019; 11. DOI: 10.3390/nu11102278.

23. Wang X, Wang H, Liu W, et al. High level of C-type natriuretic peptide induced by hyperandrogen-mediated anovulation in polycystic ovary syndrome mice. Clinical science (London, England : 1979) 2018; 132: 759–776. DOI: 10.1042/cs20171394.

24. McInnes KJ, Smith LB, Hunger NI, et al. Deletion of the androgen receptor in adipose tissue in male mice elevates retinol binding protein 4 and reveals independent effects on visceral fat mass and on glucose homeostasis. Diabetes 2012; 61: 1072-1081. 2012/03/15. DOI: 10.2337/db11-1136.

25. Cree-Green M, Torres M, Pyle L, et al. Extreme hyperandrogenism worsens metabolic but not dermatologic findings within obese adolescents with polycystic ovarian syndrome. Hormone research in paediatrics 2017; 88: 58–59. Conference Abstract. DOI: 10.1159/000481424.

26. Martínez-García M, Montes-Nieto R, Fernández-Durán E, et al. Evidence for masculinization of adipokine gene expression in visceral and subcutaneous adipose tissue of obese women with polycystic ovary syndrome (PCOS). J Clin Endocrinol Metab 2013; 98: E388–396. DOI: 10.1210/jc.2012-3414.

27. Wang J, Wu D, Guo H, et al. Hyperandrogenemia and insulin resistance: The chief culprit of polycystic ovary syndrome. Life Sci 2019: 116940. DOI: 10.1016/j.lfs.2019.116940.

28. Yang RZ, Lee MJ, Hu H, et al. Identification of omentin as a novel depot-specific adipokine in human adipose tissue: possible role in modulating insulin action. Am J Physiol Endocrinol Metab 2006; 290: E1253–1261. DOI: 10.1152/ajpendo.00572.2004.

29. Tan BK, Adya R, Farhatullah S, et al. Omentin-1, a novel adipokine, is decreased in overweight insulin-resistant women with polycystic ovary syndrome - Ex vivo and in vivo regulation of omentin-1 by insulin and glucose. Diabetes 2008; 57: 801–808. DOI: 10.2337/db07-0990.

30. Barzilai N, Wang J, Massilon D, et al. Leptin selectively decreases visceral adiposity and enhances insulin action. 1997; 100: 3105–3110.

31. Wittamer V, Franssen J-D, Vulcano M, et al. Specific Recruitment of Antigen-presenting Cells by Chemerin, a Novel Processed Ligand from Human Inflammatory Fluids. 198: 977–985.

32. Bondue B, Vosters O, de Nadai P, et al. ChemR23 Dampens Lung Inflammation and Enhances Anti-viral Immunity in a Mouse Model of Acute Viral Pneumonia. 7: e1002358.

33. Benjamin, Bondue, and, et al. Chemerin and its receptors in leukocyte trafficking, inflammation and metabolism.

34. Kim JY, Xue K, Cao M, et al. Chemerin suppresses ovarian follicular development and its potential involvement in follicular arrest in rats treated chronically with dihydrotestosterone. Endocrinology 2013; 154: 2912–2923. DOI: 10.1210/en.2013-1001.

35. Reverchon M, Cornuau M, Ramé C, et al. Chemerin inhibits IGF-1-induced progesterone and estradiol secretion in human granulosa cells. Hum Reprod 2012; 27: 1790–1800. DOI: 10.1093/humrep/des089.

36. Mangge H, Almer G, Truschnig-Wilders M, et al. Inflammation, adiponectin, obesity and cardiovascular risk. Curr Med Chem 2010; 17: 4511–4520. DOI: 10.2174/092986710794183006.

37. Jakubowicz D, Wainstein J and Homburg R. The link between polycystic ovarian syndrome and type 2 diabetes: preventive and therapeutic approach in Israel. The Israel Medical Association journal : IMAJ 2012; 14: 442-447. 2012/09/08.

38. Osibogun O, Ogunmoroti O and Michos ED. Polycystic ovary syndrome and cardiometabolic risk: Opportunities for cardiovascular disease prevention. Trends Cardiovasc Med 2019 2019/09/15. DOI: 10.1016/j.tcm.2019.08.010.

39. Tarkun I, Dikmen E, Cetinarslan B, et al. Impact of treatment with metformin on adipokines in patients with polycystic ovary syndrome. Eur Cytokine Netw 2010; 21: 272–277. DOI: 10.1684/ecn.2010.0217.

40. Ozkaya M, Cakal E, Ustun Y, et al. Effect of metformin on serum visfatin levels in patients with polycystic ovary syndrome. Fertility and sterility 2010; 93: 880–884. DOI: 10.1016/j.fertnstert.2008.10.058.

41. Li M, Yang M, Zhou X, et al. Elevated Circulating Levels of Irisin and the Effect of Metformin Treatment in Women With Polycystic Ovary Syndrome. Journal of Clinical Endocrinology & Metabolism 2015; 100: 1485–1493. DOI: 10.1210/jc.2014-2544.

42. Foda AA, Foda EA, El-Negeri MA, et al. Serum chemerin levels in Polycystic Ovary Syndrome after metformin therapy. Diabetes and Metabolic Syndrome: Clinical Research and Reviews 2019; 13: 1309-1315. Article. DOI: 10.1016/j.dsx.2019.01.050.

43. Haider DG, Schaller G, Kapiotis S, et al. The release of the adipocytokine visfatin is regulated by glucose and insulin. 2006.

44. Sharma S, Mathur DK, Paliwal V, et al. Efficacy of Metformin in the Treatment of Acne in Women with Polycystic Ovarian Syndrome: A Newer Approach to Acne Therapy. The Journal of clinical and aesthetic dermatology 2019; 12: 34–38.

